# Enhanced motor noise in an autism subtype with poor motor skills

**DOI:** 10.1101/2023.03.25.23287738

**Authors:** Veronica Mandelli, Isotta Landi, Silvia Busti Ceccarelli, Massimo Molteni, Maria Nobile, Alessandro D’Ausilio, Luciano Fadiga, Alessandro Crippa, Michael V. Lombardo

## Abstract

Early motor difficulties are a common in many, but not all, autistic individuals. These difficulties tend to be highly present in individuals carrying rare genetic mutations with high penetrance for autism. Many of these rare genetic mechanisms also cause neurophysiological dysregulation of excitation-inhibition balance (E:I). A predicted downstream consequence of E:I imbalance in motor circuitry would translate behaviorally into enhanced ‘motor noise’ – that is, increased variability in execution of motor actions. Here we tested the hypothesis that autistic individuals with the most pronounced motor difficulties would be most affected by enhanced motor noise. Unsupervised data-driven clustering on a standardized test of motor skills (n=156, age = 3-16 years) identified the presence of two robust and highly stable autism motor subtypes described by relatively intact versus highly impaired motor skills. With motor kinematics data recorded during a simple reach-to-drop task, we observed that enhanced motor noise is a specific characteristic of the autism subtype with highly impaired motor skills. Autistic individuals with poor motor skills may be differentially affected by E:I imbalance within motor circuitry.

---

The core of the autism phenotype is centered around early developmental difficulties in the domains of social-communication (SC) and restricted repetitive behavior (RRB). Despite the core SC and RRB commonalities, autistic individuals markedly vary in the domain of motor development. It has been estimated that anywhere from 34-80% of autistic individuals show some form of motor impairment and/or delay^1–5^. Motor difficulties in autism are often associated with language delay^6–8^, cognitive impairment^9–11^, poorer developmental outcomes, and reduced life quality^12–14^. Because motor difficulties may affect such a large percentage of autistic individuals, a recent debate has emerged regarding whether these issues should be added to the diagnostic criteria^1,15–18^. However, within the motor domain, the way in which motor abilities are affected in autism is quite heterogeneous. These motor issues span delays in reaching early motor milestones to severe impairments in motor coordination that prevent the individuals from accomplishing daily life tasks autonomously^13,19^. Thus, a discussion regarding global motor impairment in autism is not sufficient and may not be helpful in the context of precision medicine and personalized intervention^20,21^. An early characterization of heterogeneous motor profiles in autistic individuals is needed and may help link to other latent profiles that extend into other key domains such as language, intellectual and adaptive functioning.

Understanding motor issues in autism may also be key to honing in on biological mechanisms that affect some autistic individuals. At a genetic level, it is known that individuals with highly penetrant but rare protein truncating de novo mutations associated with autism also tend to show delays in the acquisition of early motor milestones such as age at walking^22–27^. Many of these rare highly penetrant mutations are known to converge on a final common pathway of dysregulated neurophysiological balance between excitatory and inhibitory (E:I) neuronal signaling in the brain^28,29^. Synaptic E:I imbalance can attenuate signal-to-noise ratio in neural circuitry and enhance neural noise^30^. A downstream consequence of higher neural noise in motor circuitry would lead to a behavioral prediction of enhanced motor noise - that is, increased variability when performing the same motor action repeatedly^31^. While variability is a ubiquitous and healthy feature of neuronal circuits^31–33^, enhanced neural and motor noise has been proposed to be more pronounced on-average in autism^34–37^

Here, we investigated the hypothesis of whether stratification of autism by early clinical motor profiles leads to enhanced precision in our ability to personalize explanations about enhanced motor noise to particular subsets of autistic cases with the most clear-cut atypicalities in the motor domain. As such, the current work is split into 3 objectives. First, we use an unbiased data-driven approach to identify whether there are discrete motor subtypes in autism, using early clinical profiles of motor behavior assessed with a standardized test of motor ability - the Movement Assessment Battery for Children - 2nd edition (MABC2). Second, we apply subtype labels to motor kinematics data from a simple reach-to-drop motor task^38,39^ and test whether enhanced motor noise is primarily a feature of only a subset of autistic cases with the most pronounced issues in the motor domain. Third, if enhanced motor noise due to dysregulated synaptic E:I balance in motor circuitry is a characteristic of autism or a specific autism motor subtype, we reasoned that genomic mechanisms linked to motor circuitry, autism, and developmental motor issues would converge on systems biological dysregulation of synaptic E:I mechanisms. Utilizing curated lists of genes linked to autism or developmental motor issues, alongside spatial gene expression patterning from the Allen Institute Human Brain Atlas (AHBA), we use gene expression decoding and enrichment analyses to test this hypothesis.

## Methods

### Sample and dataset characteristics

To test our first aim of stratifying autism by early clinical motor profiles, we compiled together several publicly available and in-house datasets which all use the Movement Assessment Battery for Children - 2nd edition (MABC2). The in-house dataset consisted of n=94 autistic and n=93 typically-developing (TD) children aged 3-12 years old (mean age = 7.34; SD age = 2.4), collected at the Scientific Institute IRCCS Eugenio Medea (IRCCS-MEDEA) in Italy (see Supplementary Methods for more details about the IRCCS-MEDEA dataset). Additionally, we identified another n=62 autistic, n=23 developmental coordination disorder (DCD), and n=56 TD children aged 8-16 years old (mean age = 11.2; SD age = 1.73) from the 3 datasets publicly available within National Institute of Mental Health Data Archive (NDA) in the USA (see Supplementary Methods for more details about the NDA dataset).

To evaluate our second aim of whether motor noise was enhanced specifically within an autism motor subtype, we utilized motor kinematics data from the IRCCS-MEDEA dataset. Motor kinematic data at IRCCS-MEDEA was collected with an optoelectronic system while children performed a simple upper-limb motor task^38,39^. At IRCCS-MEDEA we also collected data regarding early developmental, cognitive skills, and autism core symptoms severity from each child’s clinical portfolio. All the clinical assessments, including the diagnosis of autism, were performed by expert child-psychiatrists at IRCCS-MEDEA.

## Measures

### Movement Assessment Battery for Children - 2nd edition (MABC2)

To assess motor profiles in young children we used the Movement Assessment Battery for Children - 2nd edition (MABC2)^40^. MABC2 is a gold standard clinical test for the diagnosis of Developmental Coordination Disorder (DCD) in children aged 3 to 16 years old. However, it can be useful in assessing motor proficiency in a variety of developmental conditions, including autism. The MABC2 is composed by 3 subscales investigating specific aspects of motor coordination: the Manual Dexterity (MD) subscale, which refers to fine-motor coordination tasks; the Ball Skills (BC) subscale that includes aiming and catching activities; and the Static and Dynamic Balance (SDB) subscales that tests balance abilities. To cover a wide age range, the MABC2 is divided into 3 modules (module 1: 3-6 years; module 2: 7-10 years, module 3: 11-16 years) that assess the very same skills using age-appropriate tasks and activities. The advantages of using the MABC2 in autism is that the task’s instructions include a practical demonstration that can be followed by non-verbal autistic individuals. MABC2 has a high internal consistency (0.90) and an excellent test– retest reliability (intraclass correlation coefficient = 0.97^41^)

### Kinematic Task - Data acquisition and preprocessing

Motor kinematic data was collected during a simple reach-to-drop task previously described by Forti et al.,^39^ and Crippa et al.,^38^. The task required that the child reach to grasp a rubber ball placed in front of them, grasp it, and then drop it in a plastic container. Each child was asked to repeat this set of actions 10 times using their dominant hand. Task instructions also included a practical demonstration of the task performed by the same experimenter for all the participants. For an exhaustive description of the experimental task refers to the Supplementary Methods. Each movement was recorded using an optoelectronic system (the SMART D from BTS Bioengineering - Garbagnate Milanese, Italy). Three-dimensional kinematic data was collected by eight infrared-motion analysis cameras at 60 Hz (spatial accuracy: 0.2 mm), located four per side at 2.5 meters away from the participants. Passive markers (1 cm) were attached to the elbow, ulnar and radial surfaces of the participants’ wrists and to the hand dorsum on the fourth and fifth metacarpals (Fig. 1C). This resulted in a total of four body parts for kinematic trajectories to be recorded from. Subsequently, a dedicated software system (Smart Tracker, BTS Bioengineering - Garbagnate Milanese, Italy) was used to track and reconstruct the acquired movement by naming each single moving point recorded by the cameras in each time-frame. This allowed for frame-wise definition of a movement trajectory in 3-dimensional space coordinates (x, y, z). Data were then preprocessed in MATLAB (Mathworks - Natick, MA, USA) using a fifth-order Butterworth (8-Hz) low-pass filter. A total of n=24 autistic and n=14 TD children were excluded from the following analysis as they did not complete all 10 trials of the task.

**Figure 1:**
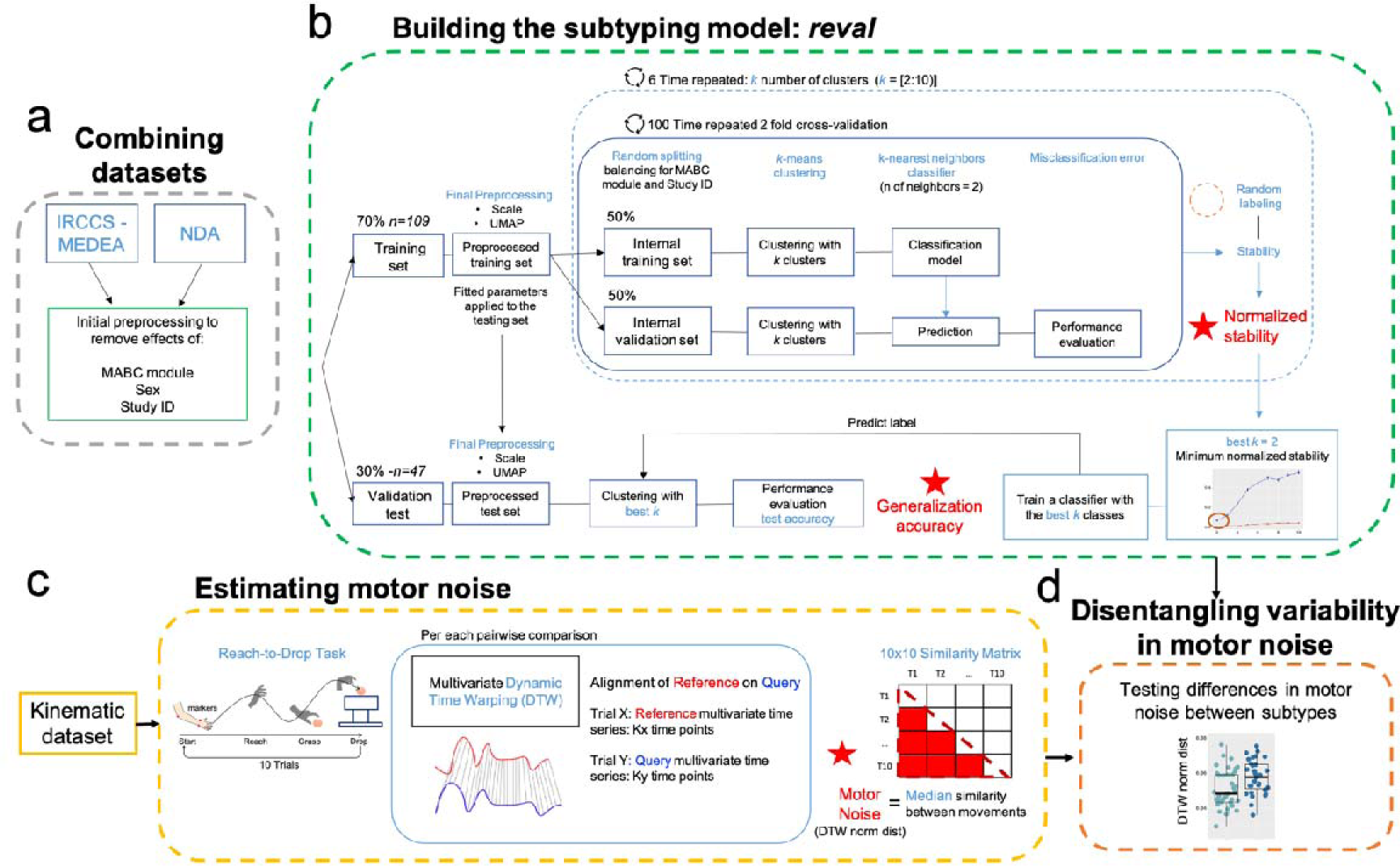
Overview of motor stratification and kinematic data analysis workflow. First, we combined the IRCCS-MEDEA and NDA datasets while doing initial preprocessing steps to remove confounding effects of originating study ID, MABC2 module, and sex (A). Panel B shows the workflow for stability-based relative clustering validation (reval) analyses that aimed to identify the optimal number of clusters (best k) that minimizes normalized stability in independent splits of the data (training and validation) and estimate the generalization accuracy of such optimal (best k) clustering solution. Panel C shows analysis workflow for how we estimated motor noise from kinematics data acquired during a simple reach-to-drop task. Here, 10 repeat trials were administered and we used multivariate dynamic time warping (DTW) to align and compare motor kinematic trajectories across the 10 repeat trials. Motor noise is operationalized as the median similarity across trials (DTW dist norm) whereby higher estimates are indicative of more motor noise (i.e., increased dissimilarity between repeat trials). Panel D shows how final hypothesis tests were done examining subtype differences in motor noise.

### Statistical analyses

For our first aim to stratify autism by MABC2 profiles, we first combined the IRCCS-MEDEA and NDA datasets to get the largest possible sample size for the stratification analysis. The final sample size was n=328 individuals aged 3 to 16 years old split into n=156 autistic (n=29 females), n=23 DCD (n=9 females) and n=149 TD (n=35 females) children. Because data originated from multiple sites (e.g. IRCCS-MEDEA and 3 other NDA datasets), we first implemented a batch correction technique to project out variance attributed to the originating study ID, MABC2 module and sex (Fig 1A). This batch correction was implemented as a linear model with each MABC2 subscale or total standardized score as the dependent variable and the originating study ID, MABC2 module, sex, and diagnosis as independent variables. Isolating the beta coefficient for the independent variable the originating study ID, MABC2 module and sex, we used this to project those features related variance before any further downstream statistical analysis. For the subsequent stratification model analysis, only the autistic subjects were utilized. For full reproducible analyses for all results presented in this work, please see https://github.com/IIT-LAND/motor_stratification_paper.

### Stratification modeling with unbiased data-driven stability-based relative clustering validation

To identify robust, stable, and reproducible subtypes based on motor profiles from the MABC2, we used our stability-based relative clustering validation approach, called *reval* (https://github.com/IIT-LAND/reval_clustering)^42^. Preprocessed MABC2 data from autistic individuals was used as input for the *reval* analysis. A total of n=156 autistic individuals were first split into *train* and *validation* sets using a 70-30 split scheme, while also balancing for originating study ID, sex, and MABC2 module. Before being entered as input features for *reval*, the three MABC2 subscales were scaled to a mean of 0 and a standard deviation of 1 (*sklearn.preprocessing.StandardScaler*) and then transformed using Uniform Manifold Approximation and Projection (UMAP) (n_neighbors = 30, min_dist = 0.0, n_components = 2, random_state = 42, metric = Euclidean). Those steps were done by fitting the models on the *train* set and applying them to both *train* and *validation* sets. Clustering and classification models were fit using k-means clustering and k-nearest neighbor classifier algorithms from the python *scikit-learn* library. To identify the optimal number of clusters via minimizing normalized stability, we used a 2-fold cross-validation scheme on the *train* dataset and searched through cluster solutions 2 through 10. The cross-validation scheme was repeated 100 times to ensure robustness. The identified optimal number of clusters was then used for clustering on the *validation* set. A classifier was then trained on the *train* set and utilized to predict the labels on the *validation* set to test the reproducibility of the cluster solution in a held-out sample. The accuracy of the classifier on the *validation* set is called *generalization accuracy* and describes how well the classification model fit to the *train* dataset can identify similar labels in the independent *validation* set (Fig 1B) (see Supplementary Methods for more information about how *reval* constructs the stratification model). While stability-based relative clustering validation in *reval* tells us about the stability of clustering solutions, it does not test whether the actual solution is indicative of true clusters. Therefore, we followed up on the *reval* analysis by using the *sigclust* library in R to test whether the observed clustering solution significantly differs from the null hypothesis that the data originates from a single multivariate Gaussian distribution^43^.

### Testing subtypes for differences in non-motor domains

Autism motor subtypes were examined for a number of phenotypic differences on autism symptom severity, autistic traits, intelligence, and age at acquisition of developmental milestones such as walking and first words. For this analysis, only individuals from the IRCCS-MEDEA dataset were analyzed, since this was the only dataset that had presence of variables measuring these non-motor domains. To measure intelligence, we utilized combined standardized scores (mean 100, SD = 15) across measures such as the Griffiths Mental Development Scales^44^, Wechsler Preschool and Primary Scale of Intelligence-III (WPPSI-III), and Wechsler Intelligence Scale for Children 3rd and 4th edition^45,46^. Autism symptom severity was measured with the Autism Diagnostic Observation Scales-2nd edition (ADOS-2)^47,48^ calibrated severity score (CSS), social affect calibrated severity score (SA), and restricted and repetitive behavior calibrated severity scores (RRB)^49^. Autistic traits were measured with the Social Responsiveness Scales (SRS-2)^50^. Age at achievement of early developmental milestones (i.e., age at independent walking, age at first words) were retrieved from the clinical history data collected by clinicians during initial examination. Hypothesis tests were conducted via Welch two-sample t-tests or Wilcoxon signed-rank tests when data significantly deviated from a Gaussian distribution (Supplementary Table 2).

### Motor kinematic analyses

Motor noise was assessed via analysis of kinematic data from a simple reach-to-drop task from the IRCCS-MEDEA dataset. Here we defined motor noise as the degree of similarity in movement trajectories between repeated trials on this task^31,51^. To estimate similarity between the 10 repeat trials of the task, we utilized multivariate dynamic time warping (DTW) implemented with the *dtw* function in the *dtw* library in *R* (distance metric: Euclidean, step pattern: Symmetric2, begin and end: close). For each individual, DTW resulted in a 10x10 similarity matrix. We defined motor noise as the median distance between trials, computed as the median in this 10x10 DTW similarity matrix. Larger values on this measure indicate higher levels of motor noise due to higher dissimilarity in movement trajectories between the 10 repeat trials (Fig 1C). Given that the movements were already segmented so that data for each trial started and ended with the starting and ending of the movement, the DTW algorithm was forced to match the first and the last timeframe while comparing trials. Moreover, to avoid any bias given by the velocity in performing the movement, the *normalized distance* output was used for the subsequent analysis which represents the difference in the trajectories normalized by the total duration of the movement. Hypothesis tests for group differences in motor noise (e.g., DTW normalized distance) were examined with ANOVA and post hoc Welch two-sample t-tests.

### Examining motor noise during feedforward and feedback phases of reaching

It is well known in the literature that reaching actions can be characterized by two phases^52–58^: 1) a first feedforward phase defined by the first deceleration peak after the max peak velocity, and 2) a subsequent feedback phase preceding the grasping of the object (Fig 4A). These phases are thought to be underpinned by distinct neurocomputational mechanisms and thus are important to separate and examine for differences between autism motor subtypes. Therefore, in addition to examining motor noise for the entire reach-to-drop action, we also examined motor noise for these specific phases of the reach action. All of the same methods used to estimate motor noise with multivariate dynamic time warping were used in these analyses. The primary difference is that the movement trajectories were segmented into feedforward and feedback phases via identifying the first deceleration peak after the max velocity peak for the reaching action. Because the two phases of the reach action can be thought of as a within-subject factor, we modeled between-group differences as potential group*phase interactions within a linear mixed effect model that treated group and phase as fixed effects and modeled random intercepts grouped by subject ID.

**Figure 2:**
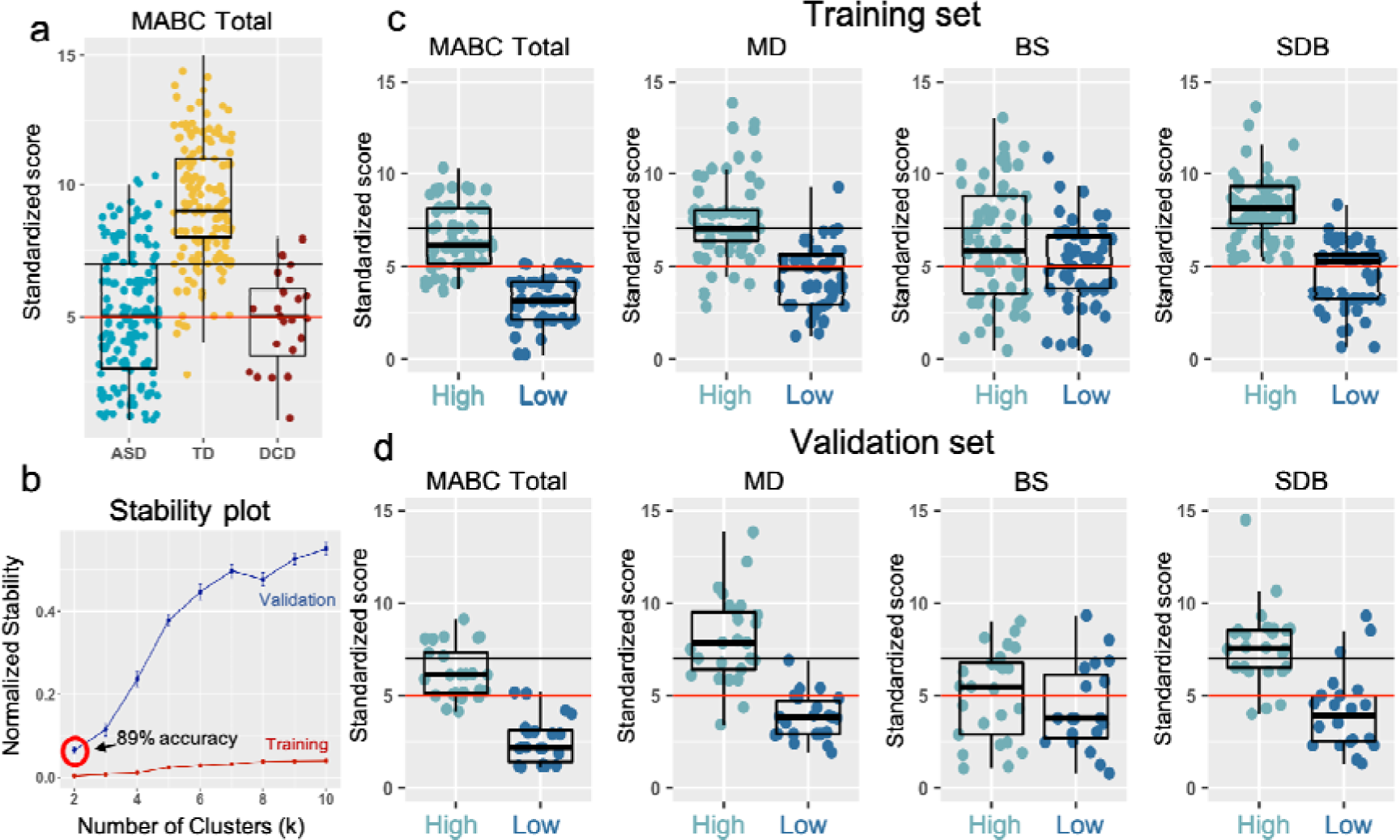
Autism motor subtypes. Panel A plots the MABC2 total standardized score for typically-developing (TD; yellow-orange), autistic (blue), and Developmental Coordination Disorder (DCD; maroon) individuals. The horizontal solid black and red lines represent cutoffs for “at risk of having a motor impairment” (solid black line) and “have a motor impairment” (solid red line) according to the MABC2 manual. With stability-based relative clustering validation (reval) analyses, the optimal clustering solution identified was k=2, indicating two subtypes that could be identified with 89% accuracy in independent data. The two subtypes (Autism High, light blue; Autism Low, dark blue) are described with respect to total MABC2 score, and the MD, BS, and SDB subscales of the MABC2 in the Training (C) and Validation (D) sets.

**Figure 3:**
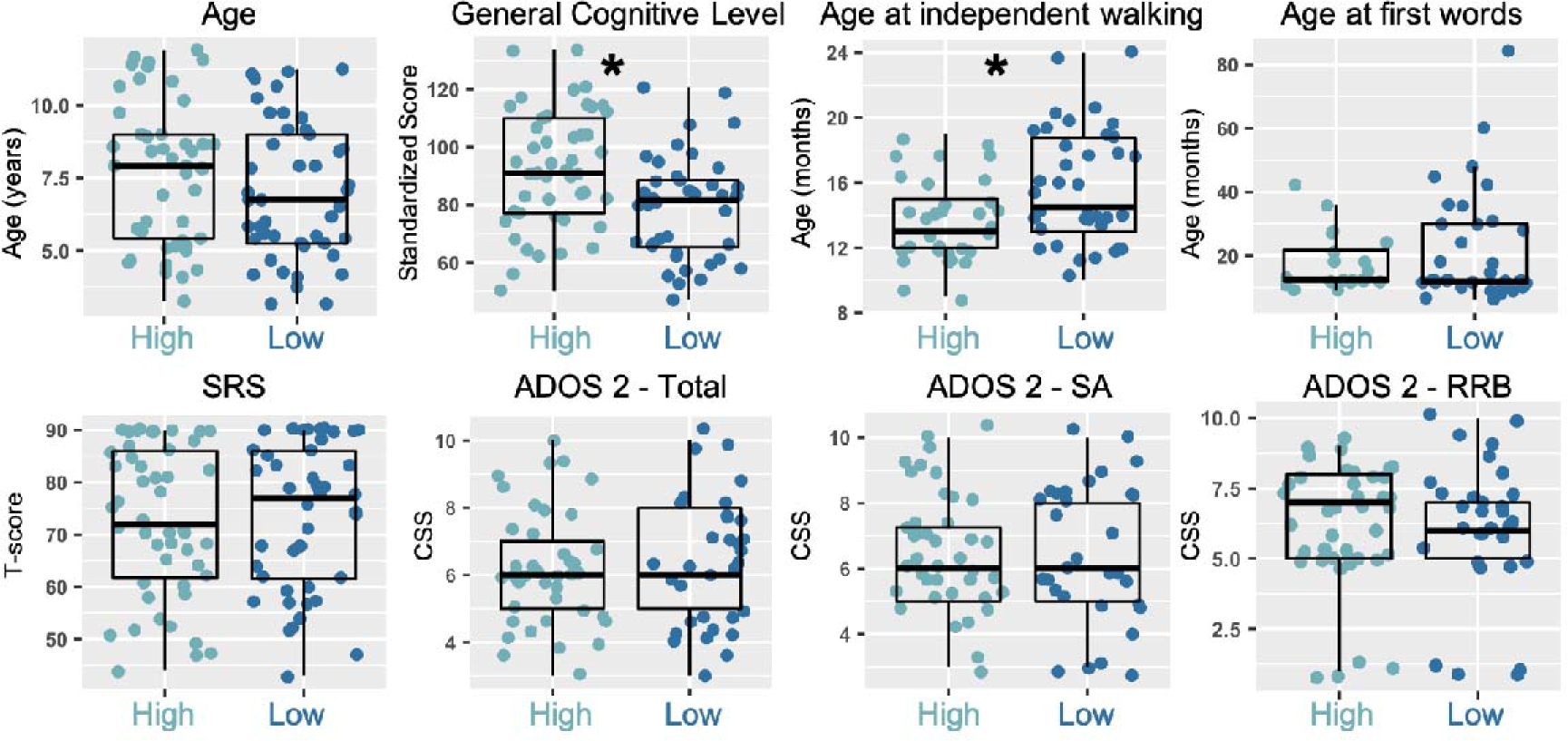
Characterization of autism motor subtypes by age, general cognitive level, early developmental milestones, and autistic traits and symptom severity. In this figure we describe subtypes (Autism High, light blue; Autism Low, dark blue) in terms of age, general cognitive level, age at acquisition of early developmental milestones (age at independent walking and first words), and autistic trait and symptom severity as measured by the SRS-2 and ADOS-2 respectively. The asterisk (*) indicates p<0.05.

**Figure 4:**
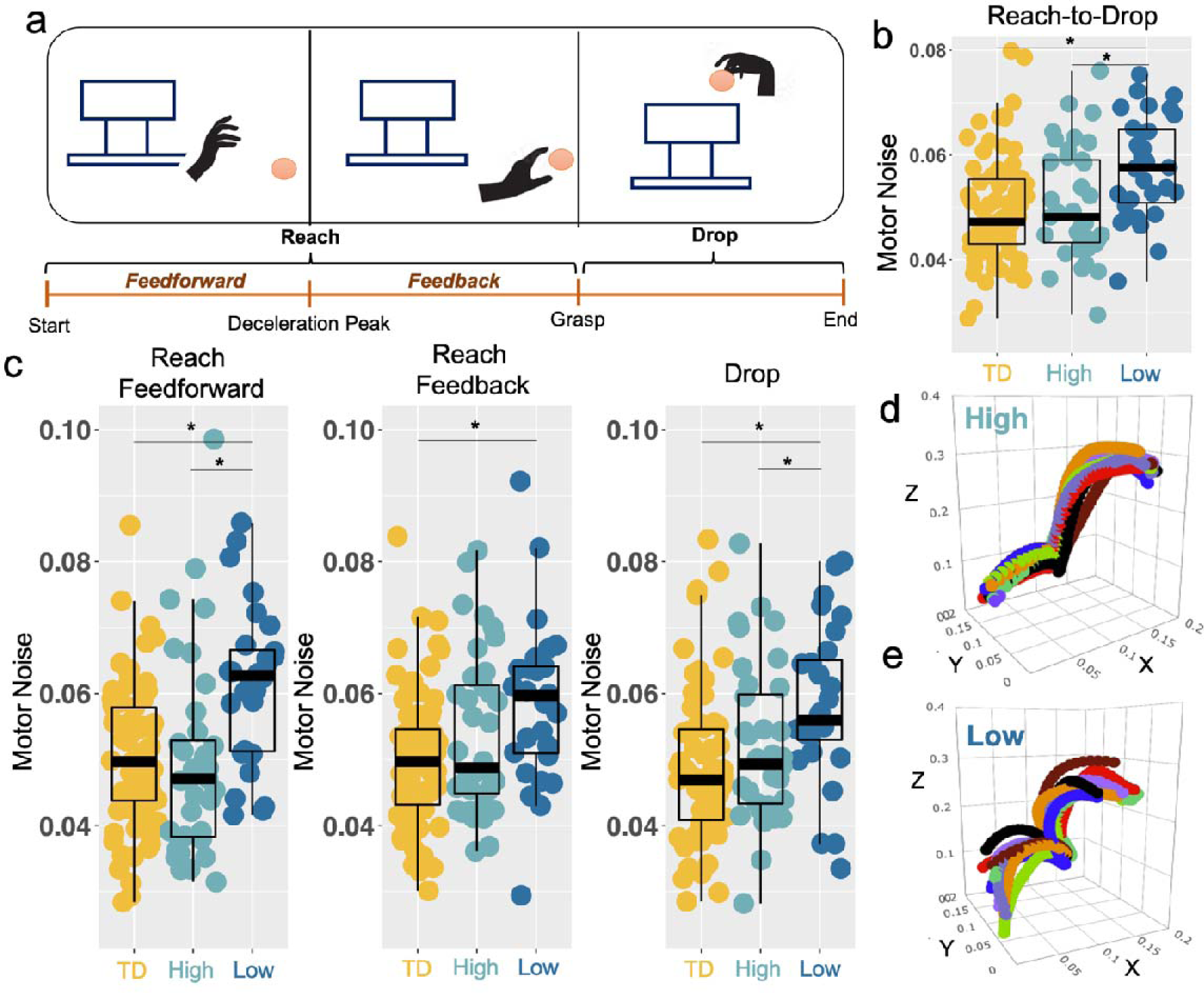
Enhanced motor noise specific to the poor motor skill autism subtype. Panel A shows the reach-to-drop task, segmented into reach and drop actions, and with the reach action split into feedforward and feedback phases according to the deceleration peak. Panel B shows group differences (TD, yellow-orange; Autism High, light blue; Autism Low, dark blue) when motor noise is measured across the entire reach and drop actions, motor noise for each subject is operationalized as the median normalized distance between 10-trials movement trajectories computed with multivariate Dynamic Time Warping. Panel C shows group differences when the task is split into reach and drop actions and with the reach action split into feedforward and feedback phases. Panels D and E show example trajectories (one line per trial) in the original 3D space (X, Y, Z) for one body part (lateral wrist) for randomly selected ‘High’ and ‘Low’ autistic individuals, in order to visually show how motor noise can be understood as higher levels of variability across repeat motor actions. The asterisk (*) indicates p<0.05.

### Gene expression decoding and enrichment analyses

The third aim of this work was to test whether genomic mechanisms linked specifically to motor circuitry, autism, and developmental motor issues would converge on systems biological dysregulation of synaptic E:I mechanisms. To achieve these goals, we first utilized gene expression decoding analysis to isolate genes that spatially express in patterns that are highly similar to topology of macroscale networks isolated in previous resting state functional connectivity studies. For motor circuitry, we use a resting state fMRI (rsfMRI) connectivity map of well-known motor circuitry that covers motor and premotor cortex as well as cerebellar motor areas (Fig 5A). In contrast to motor circuitry, we also utilized all other non-motor related macroscale rsfMRI network maps (Supplementary Figure 1) defined from previous work utilizing independent components analysis to identify major well-known rsfMRI networks^59^. These networks comprise a range of cortical, subcortical and cerebellar networks, but exclude motor circuitry. Gene expression decoding was implemented via Neurosynth^60^ to identify genes that are statistically similar in their expression profile in a consistent manner across all six donor brains within the Allen Institute Human Brain Atlas (AHBA)^61^. The analysis first utilizes a linear model to compute the similarity between the motor connectivity map and spatial patterns of gene expression for each of the six brains in the AHBA dataset. The slopes of these subject-specific linear models encode how similar each gene’s spatial expression pattern is with our motor circuit connectivity map. These slopes were then subjected to a one-sample t-test to identify genes whose spatial expression patterns are consistently of high similarity across the donor brains to the motor connectivity map. This analysis was run using the full patterning across cortical, subcortical, and cerebellar areas. The resulting list of genes was then thresholded for multiple comparisons and only the genes surviving FDR q<0.05 and which had a positive t-statistic value were considered further.

**Figure 5:**
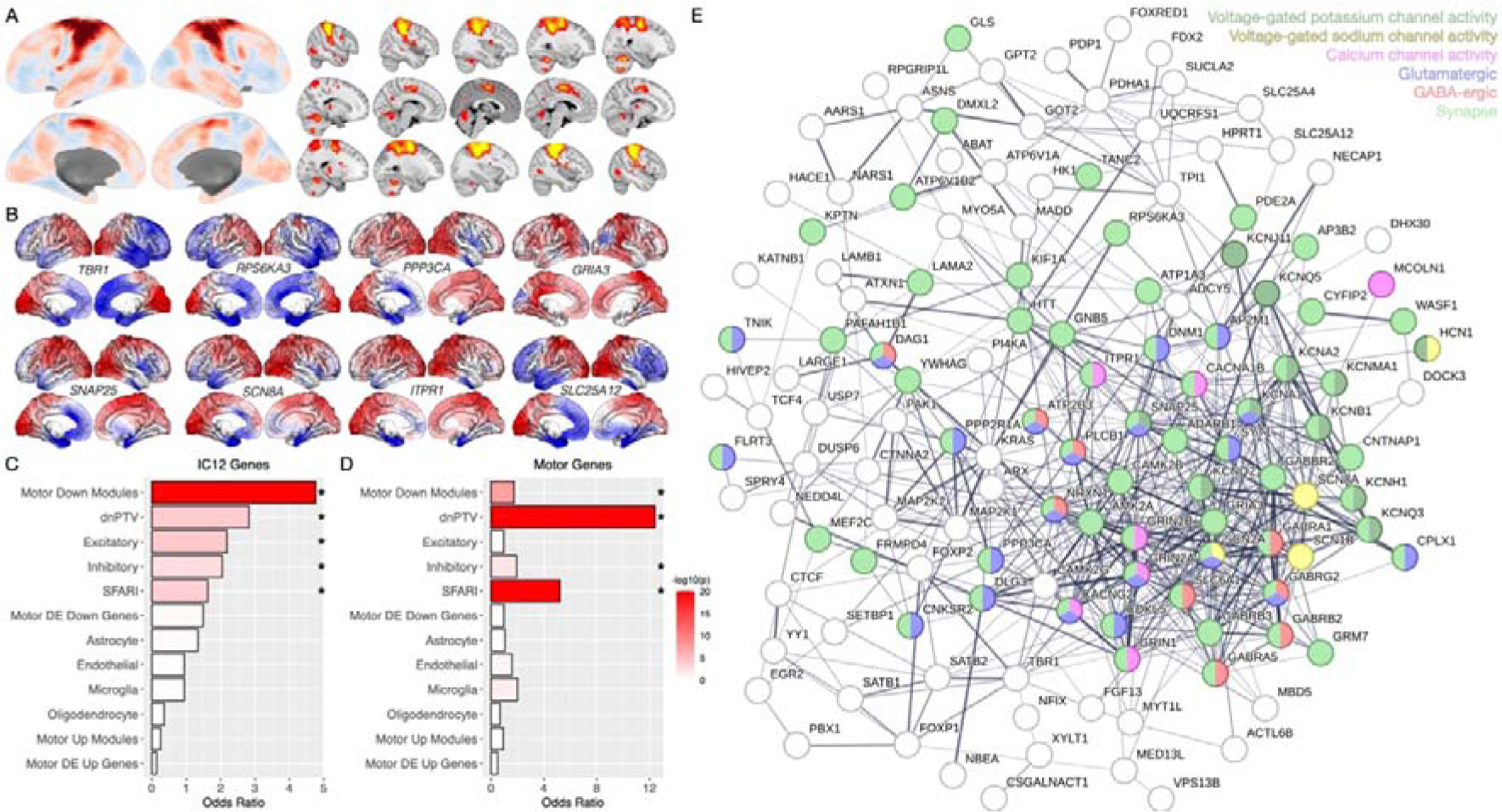
Genes associated with autism and developmental motor issues are specifically enriched for genes highly expressed within motor circuitry and highlight E:I imbalance mechanisms. Panel A shows cortical, subcortical and cerebellar motor circuitry isolated with resting state fMRI connectivity analysis (taken from the IC12 rsfMRI connectivity map from Bertelsen et al., 2021). Gene expression decoding analysis was implemented on this motor circuit map to identify the subset of genes showing evidence for highly similar spatial expression patterns to this motor circuit. Examples are shown in panel B of 8 genes that show strong spatial expression similarity to motor circuitry and which are also autism-associated and linked to developmental motor issues. Genes linked to IC12 motor circuitry (C) or developmental motor issues (D) are enriched for similar profiles of autism-associated genomic mechanisms, as shown in panels C and D respectively. These plots show the enrichment odds ratio for each of the 12 autism-associated gene lists. The asterisk indicates significant enrichments that pass FDR q<0.05. Increasingly red color indicates increasing statistical significance of the enrichment test, as indicated by the -log10 p-value. Panel E shows a network plot of protein-protein interactions between genes that are associated with autism and developmental motor issues and are highly expressed specifically within motor circuitry (IC12). Colors for some nodes indicate involvement at the synapse, glutamatergic or GABA-ergic transmission, or with ion channel activity.

After identifying genes that spatially express in a highly similar manner to motor circuitry, we next used gene set enrichment analysis to test whether motor circuitry-relevant genes are enriched for genes known to be associated with autism or developmental motor issues. For the autism-associated gene lists, we utilized an array of gene lists from past autism genomics work such as genes from the expert curated SFARI Gene database (list labeled in Fig 5C-D as SFARI) (https://gene.sfari.org/; release 01-23-2023), rare de-novo protein truncating variants associated with autism (list labeled in Fig 5C-D as dnPTV)^25^, genes and co-expression modules differentially expressed in autism from motor (BA3/1/2/5) and premotor (BA4/6) regions (lists labeled in Fig 5C-D as Motor Up or Down Modules and Motor DE Up or Down Genes)^62^, and genes differentially expressed from neuronal and non-neuronal cell types (lists labeled in Fig 5C-D as Excitatory, Inhibitory, Astrocyte, Microglia, Endothelial)^63^. For genes associated with developmental motor issues, we combined gene lists curated in the Monarch database (https://monarchinitiative.org)64 for the phenotypes of ‘delayed ability to walk’ (HP:0031936), ‘delayed gross motor development’ (HP:0002194), ‘hypotonia’ (HP:0001252), ‘motor delay’ (HP:0001270), and ‘poor gross motor coordination’ (HP:0007015). All enrichment tests computed with custom code written in R that computes enrichment odds ratios and p-values based on hypergeometric tests. The background total for these enrichment tests was set to 16,796, which is the total number of genes considered in prior work as robustly expressed in brain tissue^62^. Only tests that survived FDR q<0.05 were interpreted further as statistically significant enrichments.

Genes that were identified as associated with autism and developmental motor issues were tested for overlap with genes highly expressed in either motor circuitry, cortical and subcortical networks, and a cerebellar network (Supplementary Figure 1). Overlapping autism- and developmental motor issue and motor circuitry genes were used in a protein-protein interaction analysis, implemented in STRING (https://string-db.org/) with the default settings of the full STRING network, usage of all interaction sources, a minimum required interaction score of 0.4, and restricted to only the query proteins. Gene ontology molecular function and cellular compartment enrichment terms were identified from this analysis that showcased involvement of different synaptic compartments and ion channel molecular functions.

## Results

### Identification of autism motor subtypes with data-driven clustering

Our primary goal of this work was to examine heterogeneity in early motor profiles in autism. However, before diving into analyses that examine such heterogeneity, it is important to first describe how autism can be characterized as a whole via a case-control analysis relative to both a TD group and also a non-autistic comparison group with significant motor impairment (DCD). Large group differences are apparent in MABC2 total standardized score (*F* = 148, *p* =2.01e-46), which can be described as lower scores in autism compared to TD (*t(302.76)* = -16.37, *p* =1.42e-43, *Cohen’s d* = 1.87), but no case-control differences when autism is compared to DCD (*t(34.35)* = 0.29, *p* = 0.77, *Cohen’s d* = 0.05) (Fig 2A).

These case-control analyses may indicate that very prominent motor impairments are a key characteristic of autism as a whole. However, before making this interpretation, it is important to analyze whether the autism group could be split into robust, stable, and reproducible subtypes. Evidence supporting the notion that autism can be split into subtypes may help refine the precision of our interpretations about motor skills for specific types of autistic individuals. To test this, we analyzed multivariate clinical motor profiles from the MABC2 for evidence of robust, stable, and reproducible autism subtypes with stability-based relative clustering validation (*reval*) analysis^42^. Our analysis revealed the existence of two autism motor subtypes with high generalization accuracy (89%) in independent data (Fig 2B). These subtypes showed evidence of separated clusters with tests against the null hypothesis that they originated from one multivariate Gaussian distribution (SigClust *p* =9.999e-05). The subtypes can be described as relatively ‘High’ (55%) versus ‘Low’ (44%) levels of motor proficiency, with subscales such as Manual Dexterity (MD) (*Cohen’s d* for the training set = 1.55, validation set = 2.18) and Static and Dynamic Balance (SDB) (*Cohen’s d* for the training set = 2.06, validation set = 1.73) showing the largest differentiation between the subtypes (Fig 2C and 2D), whereas much less differentiation exists between subtypes in the Ball Skills (BS) subscale (*Cohen’s d* for the training set = 0.43, validation set = 0.54). Notably, the subtypes are identified with high generalization accuracy without visible evidence of hard cutoffs (Fig 2C-D). However, it is apparent when data is plotted with traditional MABC2 cutoffs for motor impairment that the relatively ‘Low’ subtype scores at or below this threshold, while the relatively ‘High’ subtype is largely above this cutoff for a majority of the autistic individuals (Fig 2C-D). With robust and stable subtype labels known, we can re-evaluate our model of group differences considering the autism subtypes along with TD and DCD. With the model comparison Akaike Information Criterion (AIC) statistic, this model is significantly better at explaining variance in total MABC2 standardized scores (ΔAIC = 137). We can also describe how the ‘Low’ and ‘High’ subtypes compare relative to the TD and DCD comparison groups. The ‘Low’ subtype is more than 3 SD below the TD group (*t(209.02)* = -26.98, *p* = 4.43e-70, *Cohen’s d* = 3.22), and is also more impaired than the DCD group, even though the effect size is much less pronounced (*t(28.32)* = -2.99, *p* = 0.005, *Cohen’s d* = 0.89). In contrast, the ‘High’ group still shows lower on-average scores compared to TD (*t(255.93)* = -11.81, *p* = 2.19e-25, *Cohen’s d* = 1.46), but they show higher scores than DCD (*t(30.17)*= 4.94, *p* = 2.69e-5, *Cohen’s d* = 1.3). This indicates that the overall lack of case-control difference between autism and DCD described earlier was driven primarily because of the ‘Low’ autism subtype.

We next tested for differences between the subtypes on skills outside of the motor domain. No significant difference exists in age between subtypes (*Wilcoxon Z* = 1270, *p* = 0.2). Given literature showing that motor impairments tend to also be associated with other impairments in cognitive skills as well as achievement of early developmental milestones, we discovered that the relatively ‘Low’ motor skill autism subtype shows significantly lower scores in general cognitive ability (*t(85.96)* = 3.3, *p* = 0.001, *Cohen’s d* = 0.7) and age at independent walking (*Wilcoxon Z* = 417, *p* = 0.015, *Cohen’s d* = 0.67). However, no significant difference was apparent for age at first words (*Wilcoxon Z* = 343.5, *p* = 0.95). Similarly, no significant differences were apparent for ADOS measures of symptom severity or autistic traits measured by the SRS (Fig 3 and Supplementary Table 2).

### Autism motor subtypes are differentiated by motor noise

So far, we have demonstrated that autism can be separated into at least two subtypes by clinical motor profiles. Our second aim of this work was to test whether such subtypes are highly differentiated in terms of motor noise. Using motor kinematics data from the IRCCS-MEDEA dataset during a simple reach-to-drop task, we characterized motor noise as the degree of similarity between multivariate movement trajectories during 10 repeat executions of those actions. Higher levels of motor noise are indicated by higher DTW normalized distance. We find highly significant differences between groups in motor noise (*F* = 8.7, *p* = 2.5e-4), driven by enhanced motor noise in the relatively ‘Low’ subtype compared to the relatively ‘High’ subtype (*t(66.87)* = -3.2, *p* = 0.002, *Cohen’s d* = 0.77) and compared to TD children (*t(60.11)* = -4.1, *p =* 1.33e-4, *Cohen’s d* = 0.85). In contrast there was no difference between the relatively ‘High’ subtype and TD children (*t(71)* = -0.4, *p* = 0.66, *Cohen’s d* = 0.08) (Fig 4B). This result reflects poorer precision, and thus higher variability, in repeat executions of the action that is specific to the relatively ‘Low’ subtype. Illustrative examples of this effect can be seen in Fig 4D-E, whereby it is visually evident that the individual in the ‘High’ subtype shows very similar trajectories for each of the 10 repeat executions, while the example individual in the ‘Low’ subtype shows much more variable trajectories for each execution.

### Enhanced motor noise during the feedforward phase of reaching

The reaching component of our task can be broken down into two functionally separable components - feedforward and feedback phases. The transition between these phases is demarcated by the end-point (i.e., wrist) transport deceleration peak^57^. Therefore, we re-examined motor noise when the data is split into these two phases. Linear mixed effect modeling was able to identify a significant group*phase interaction (*F* =3.5, *p* = 0.03) which is indicative of group differences in motor noise that are dependent on the phase (feedforward or feedback). Follow-up tests showed that the autism subtypes are highly differentiated during the feedforward phase (*t(61.87)* = -3.56, *p* = 7e-4, *Cohen’s d* = 0.87), but were not different during the feedback phase *(t(58.05)* = -1.55, *p* = 0.12, *Cohen’s d* = 0.39) (Fig 4C). Examination of motor noise for the drop action also indicated that the ‘Low’ subtype showed more motor noise than the ‘High’ subtype (*t(57.87)* = -2.05, *p* = 0.045, *Cohen’s d* = 0.51) (Fig 4C). Comparing the autism subtypes to TD children, we observed a similar expression of motor noise in the relatively “High” subtypes in the both feedforward and feedback phase and in the drop action, while the motor noise of the relatively “Low” subtype is always higher with respect to TD children (Supplementary Table 4). Overall, these results demonstrate that enhanced motor noise is a specific characteristic of the relatively ‘Low’ motor skill autism subtype and that this effect could be most pronounced within neural circuitry that supports computations critical for feedforward processing.

### Genomic mechanisms behind enhanced motor noise via synaptic E:I imbalance

In the final set of analyses, we used genomics datasets and approaches to isolate candidate mechanisms that may underpin poor motor skills and enhanced motor noise via synaptic E:I imbalance. To test this hypothesis, we first isolated a subset of genes that spatially express in patterns that are highly similar to motor circuitry, via gene expression decoding analysis on an input map of whole-brain motor circuitry spanning motor and premotor cortex as well as cerebellar motor regions^59^ (Fig 5A-B; Supplementary Table 5). These motor circuit genes are highly enriched with autism-associated genes - particularly, genes that harbor high-penetrant mutations linked to autism (e.g., SFARI, dnPTV), genes from autism-downregulated co-expression modules from motor and premotor cortex, and genes differentially expressed in autism within excitatory and inhibitory neuronal cell types (Fig 5C; Supplementary Table 6). This result suggests that a number of important autism-associated genomic mechanisms can have prominent impact on motor circuitry. A very similar profile of autism-associated enrichment was observed with genes linked to developmental motor issues (Fig 5D; Supplementary Table 6), with the strongest enrichment for rare de-novo protein truncating (dnPTV) genes. Genes that are both autism-associated and associated to developmental motor issues are significantly enriched with that are genes highly expressed in motor circuitry (OR = 1.64, p = 0.002). In contrast, no significant enrichment can be identified with a collection of all other genes highly expressed across a range of other cortical and subcortical (OR = 2.49, p = 0.44) or a cerebellum-only network (OR = 1.06, p = 0.99). These results suggest that genes associated to both autism and developmental motor issues overlap in a specific-manner with genes highly expressed in motor circuitry compared to the rest of the brain. With a protein-protein interaction analysis, we also identified that motor circuit-relevant genes that are also associated with autism and developmental motor issues strongly interact at the protein level (observed edges = 591, expected edges = 177, *p* < 1e-16), and is characterized by key synaptic E:I-relevant enrichments affecting glutamatergic and GABA-ergic synapses (e.g., *GRIN2A, GRIN2B, GRIN1, GABRB2, GABRG2, GABRA1, GABRA5, NRXN1, SNAP25, DLG3*) as well as ion channel activity (e.g., calcium channels, voltage-gated sodium and potassium channels; SCN2A, SCN1B, SCN8A, *KCNQ2, KCNQ3, KCNQ5, KCNA1, KCNA2, KCNMA1, KCNJ11, KCNB1, KCNH1, CACNA1B*, *CACNG2, HCN1*) (Fig 5E). All together, these results highlight that genomic mechanisms highly expressed in motor circuitry and associated with autism and developmental motor issues can be described by a highly interacting set of proteins that prominently affects synaptic E:I balance.

## Discussion

In this work we aimed to study heterogeneity in early motor ability in autism. Past work has indicated that motor issues are a very prominent feature of autism and that it could be potentially important to consider adding this domain to the diagnostic criteria in the future^1,16^. Congruent with these ideas, if one were to simply use cut-off scores on the MABC2, we would find that a large majority of autistic individuals in our sample (74%) show medium to severe motor impairments, while only 26% possess motor skills in line with age-expected norms. However, this way of analyzing the data does not rigorously test whether autism is indeed a single group or a collection of different subtypes in the motor domain. Our work shows first and foremost that when considering motor ability in autism, the data do not conform to a single unitary group. Rather, autism can be split in an unbiased and data-driven manner into two subtypes – relatively ‘High’ versus ‘Low’ groups. These ‘High’ versus ‘Low’ subtype labels are intended as descriptive terms referencing the scores of MABC2 test and are not meant to be interpreted in relation to functioning level of each subtype. While the autism group shows on-average lower standardized scores on the MABC2, this lower level of motor ability is clearly driven by the ‘Low’ subtype, which considerably drives down the overall average score of the autism group. However, even the relatively ‘High’ subtype identified here is still on-average lower than the TD group (*Cohen’s d* = 1.46). Nevertheless, this relatively ‘High’ group is still higher than a non-autistic group of individuals with very pronounced motor impairments (e.g., the DCD group; *Cohen’s d* = 1.3). Finally, the percentages of individuals in these two subtypes (Low = 44%; High = 55%) do not easily conform to the percentages seen when one uses standardized cutoff scores on the MABC2 (e.g., 74% vs 26%). This result illustrates the need to characterize autistic individuals not only by where they stand relative to TD norms, but also with regards to how they are grouped within the autism population^65^.

After identifying heterogeneity in early motor skills in autism, we next found that the relatively ‘Low’ autism motor subtype could be characterized by enhanced motor noise during a simple reach-to-drop task where fine-grained motor kinematics were measured. Motor noise is defined as the degree of variability in repeat motor actions^31^ and is thought to be a downstream consequence of neural noise within motor circuitry^30^. The concept of neural noise can be linked to long-standing ideas in autism research such as the E:I imbalance theory^28,29^. It is known that many highly penetrant rare genetic mutations associated with autism also highly dysregulate E:I balance^28,29^ and these types of genetic mutations are often associated with delays in acquiring early motor milestones^22–27^. Enhanced motor noise specific to the relatively ‘Low’ autism motor subtype may be revealing of very different neurobiological mechanisms linked to synaptic E:I imbalance in motor circuitry. Genomics analyses supported these inferences, as we identified a convergence of genes highly expressed within motor circuitry that are also associated with autism and developmental motor issues. Highlighting the specific importance of canonical motor circuitry, in other analyses we found no significant enrichment with genes highly expressed across many other non-motor related cortical and subcortical networks or a cerebellum-only network. Autism- and developmental motor issue genes that are highly expressed in motor circuitry also highly interact at the protein-level and are integrally involved with mechanisms of direct relevance to synaptic E:I balance (e.g., glutamatergic and GABA-ergic synapses, ion channel activity). These results point to the possible interpretation that atypical genomic mechanisms linked to autism and poor motor development act specifically on motor circuitry to disrupt synaptic E:I balance and could help to explain why enhanced motor noise is a key characteristic of the poor motor skill autism subtype. With regards to how these insights could help drive future work, we suggest that new work could utilize our motor stratification model to examine how these motor subtypes might be different with respect to biomarkers relevant to E:I imbalance in neuroimaging data^66^, particularly with respect to cortical motor circuitry. If E:I imbalance is a key neurobiological issue in the ‘Low’ motor subtype, it may be important to utilize our stratification model in clinical trials that target key E:I mechanisms ^67–69^ and their effects on motor circuitry. Other future work could examine how rare variant or polygenic genomic architecture may affect motor circuitry in a differential manner in phenotypically-defined autism subtypes where motor skills are the central differentiating factor.

While motor noise highly differentiated the subtypes for trajectories analyzed across the entire reach-to-drop task, we also discovered that enhanced motor noise in the ‘Low’ subtype may be most pronounced for the feedforward phase of the initial reach action. This result is consistent with previous studies that provided evidence for alterations in the feedforward-based phase^34,70,71^. This result is also important with respect to the hypothesized different neurocomputational mechanisms that underlie feedforward versus feedback motor control. Motor control is based on the integration of feedforward action planning and feedback-based control processes. Feedforward processing derive from internal representations of the action that specify a relatively coarse motor output prior to its initiation, while feedback processes fine-tune the motor output on the fly, relying on sensory feedback and often applying corrective adjustments^72^. Action representations, as well as the neural machinery required to adapt them to incoming sensory information, are believed to rely on the cerebellum^53,73,74^. Altered cerebellar function during development might play a key role in contributing to both motor and non-motor alteration in autism^75^. Altered feedforward and feedback mechanisms are also associated with the severity of communication impairments in autism and could potentially reflect the respective contributions of the anterior and posterior cerebellum^76^. Reduced motor noise during the feedforward phase for the ‘High’ autism motor subtype suggests that this subgroup, rather than having better feedback-based correction abilities, are characterized by relatively more preserved representation of actions.

In contrast to the sharp differences between autism motor subtypes in terms of general cognitive ability, acquisition of early motor milestones, and motor noise, these subtypes were not highly differentiated in terms of age, autistic traits, or autism symptom severity. This lack of differentiation in autistic traits and core autism symptom severity is important because it potentially underscores the orthogonal nature of motor versus core diagnostic features of autism (e.g., SC and RRB domains). An emerging literature is building indicating that the single diagnostic label of autism is not enough for understanding clinical and biologically important features within autistic individuals^77,78^. Rather than looking to core SC and RRB features, it seems that a constellation of related features that do not represent the core features of autism, such as motor, language, intellectual, and adaptive functioning, may better separate out important clinical and biological distinctions within the autism population. Supporting this statement, there is evidence showing that motor difficulties in autism tend to highly co-occur with language delay^6–8^, cognitive impairment^9–11^, poorer developmental outcomes, and reduced life quality^12,14^. Similarly, individuals with very poor early language outcome tend to also have extensive issues in motor, non-verbal cognitive ability, and adaptive functioning, and also have very different structural and functional neural mechanisms underpinning their difficulties^79–82^. A theoretical advance forward for the field would be to put together these findings under a model that supports the fact that a primary split in the autism population should be between individuals with very pronounced issues in this constellation of non-core features in motor, language, intellectual, and adaptive functioning. We have proposed such a theory and have provided initial evidence in support of this subtyping model^65^. The current work identifying 2 discrete subtypes in the motor domain and which have differences extending into other domains like general cognitive ability alongside potential differences in underlying neural mechanisms matches the predictions of our model and provides further empirical support for it.

In conclusion, we have shown evidence that autism can be split into two subtypes based on early clinical motor profiles measured by the MABC2. These subtypes show other differences in general cognitive ability, acquisition of early motor milestones, and motor noise. However, they are not different with regards to autistic symptomatology. Our findings fit with general findings that autism can be stratified into robust/stable and discrete subtypes and that such subtypes including motor issues may be very relevant to the larger scope of subtypes that share issues across language, intellectual, and adaptive functioning but are likely orthogonal to issues within the core autism domains of SC and RRB.

## Data Availability

Publicly available data re-analyzed for this work can be found on the National Institute of Mental Health Data Archive (NDA). All other in-house data from the present study are available upon reasonable request to the authors.

## Acknowledgments

This project has received funding from the European Research Council (ERC) under the European Union’s Horizon 2020 research and innovation programme under grant agreement No 755816 (AUTISMS) (ERC Starting Grant to MVL). Data collection and research from IRCCS-MEDEA has been funded by grants from the Italian Ministry of Health to AC (Ricerca Finalizzata GR-2011-02348929; Ricerca Corrente 2023, “Progetto Mosaico”)”

## Author Contributions

Conceptualization: VM, AC, MVL. Methodology: MVL, IL, VM, AC, ADA, LF. Software: IL, VM, MVL. Formal analysis: MVL, VM. Investigation: MVL, VM, SBC, MM, MN, AC. Data curation: MVL, VM, AC. Writing - original draft preparation: MVL, VM, AC, ADA, LF. Writing - review and editing: MVL, VM. Visualization: MVL, VM. Supervision: MVL, AC. Project administration: MVL, AC. Funding acquisition: MVL, AC.

---

**Supplementary Figure 1:**
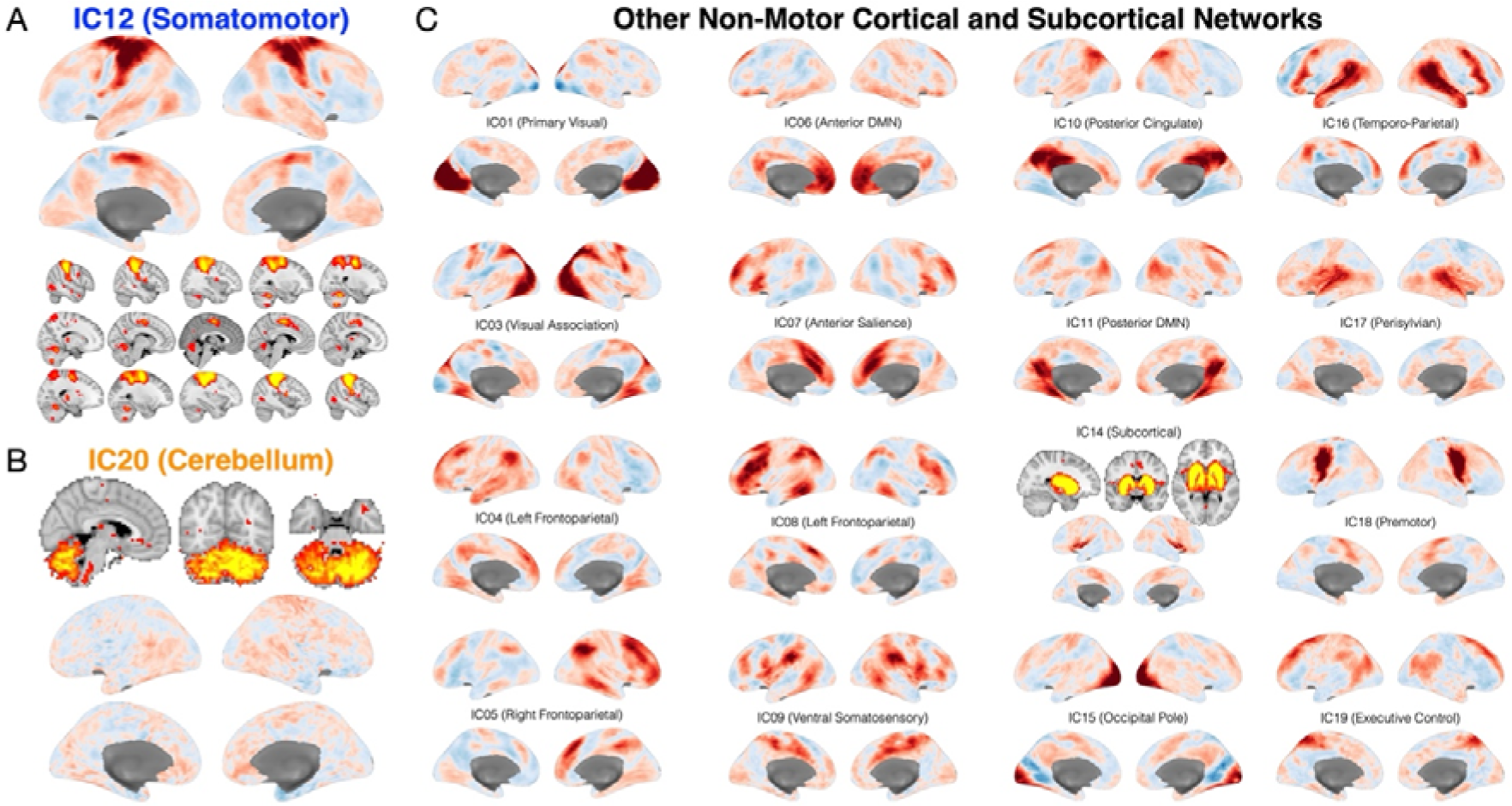
All rsfMRI networks analyzed in gene expression decoding analyses. These networks were isolated from a group independent components analysis (ICA) previously reported in our work^59^. The three panels in this figure reflect the networks that were analyzed as a set in the gene expression decoding analysis and subsequent enrichment analysis. First, a gene expression decoding analysis was done for each IC map. Then the subsequent genes surviving multiple comparison correction at FDR q<0.05 were selected and used in an enrichment analysis to test for significant overlap with a gene set associated with both autism and developmental motor issues. Panel A shows the primary component reflecting motor circuitry (IC12). Panel B shows the cerebellar network (IC20). Panel C shows all other non-motor related cortical and subcortical networks. Since there are many ICs in this collection of other non-motor related cortical and subcortical networks, we first concatenated together all genes from the gene expression decoding analyses of each IC map, and then ran the final enrichment test on this large concatenated list of genes from all of these networks combined.

