## Supplementary Methods for "Enhanced motor noise in an autism subtype with poor motor skills"

***Datasets***

***IRCCS-MEDEA dataset***

Description of the IRCCS-EM dataset can be found elsewhere in previously published work using this dataset[1]. The dataset used in this work included 94 autistic and 93 typically developing children aged 3 to 12 years old. The autistic individuals had been recruited while undergoing either a clinical assessment (diagnostic or follow-up) or comprehensive rehabilitation program at the Child Psychopathology Unit of Scientific Institute, IRCCS Eugenio Medea (Bosisio Parini, Italy), where all the testing occurred. Work utilizing this dataset was approved by "Comitato Etico IRCCS-E. Medea – Sezione Scientifica Associazione La Nostra Famiglia"; the parents provided written informed consent according to the Declaration of Helsinki. The experimental procedure included the administration of the Movement Assessment Battery for Children 2nd Edition (MABC) and the performance of 10 trials of a Reach-to-Drop motor task[1–3]. Moreover, additional information was retrieved for from each autistic individual's closest clinical record. They included information about: early development (i.e. the age at independent walk and the age of the first words), cognitive abilities (tested with the Griffith Mental Developmental Scales - 3rd Edition (GMDS-3), the Wechsler Preschool and Primary Scale of Intelligence- III (WPPSI-III), the Wechsler Intelligence Scale for Children 3rd and 4th edition) and autism core symptoms severity (using the Calibrated Severity Score (CSS), for the Total, the Social Affect (SA) and the Restricted and Repetitive Behavior (RRB) domains of the Autism Diagnostic Observation Scales- 2nd edition (ADOS-2)[4] and the Social Responsiveness Scales - SRS). It was not possible to retrieve that information for all the 94 autistic individuals, Supp Table 3 provides a description of the sample size for each of the investigated features. 24 autistic individuals did not complete all the 10 trials of the Reach-to-Drop motor task; thus they were excluded from the second set of the analysis.

***NDA dataset***

The National Institute of Mental Health Data Archive - NDA ( https://nda.nih.gov) dataset was downloaded in March 2020. It is composed of subjects identified as having completed an assessment using the Movement Assessment Battery for Children (MABC) - 2nd Edition. Available data originated from 3 different NDA collections: 2566 n=38, 2254 n=88, 2799 n=15. Supplementary table 1 reports the NDAR GUI of the subjects included in this study, along with their collection IDs. A fourth data collection (collection ID = 2093) was identified yet excluded for the unusual data distribution (i.e., most of the individuals included had a score at the lower end of the scale). The MABC data from the were filtered to only include individuals between 3 and 16 years of age. Duplicate data were identified and dropped. Finally, if more than one ABC subscale domain was missing, the subject was dropped from the analysis.

***Measures***

***Kinematic Task - Data acquisition and preprocessing***

The Reach-to-Drop motor task has been previously used and described in the following papers[2,3,5]. We briefly describe the essential part of the task useful for understanding the current study. Each trial started with the child's hand resting in a set position, at a distance equal to the 80% of each child's forearm length from the ball support. The child was asked to complete 2 subsequential movements: 1) grasp a rubber ball (6-cm diameter), placed over a small support, and 2) drop it in a plastic "castle" with a 7 cm diameter hole on the top. The "castle" was a see-through square box (21 cm high and 20 cm wide) large enough not to require fine movements while dropping the ball. Task instructions were provided by a practical demonstration with no verbal cues so that also non-verbal and minimally verbal autistic individuals could take part in the experiment. The same experimenter was present for all the session in order to avoid any possible confounds due to the practical demonstration.

Practice trials, the number of which varied individually, were given to participants before recording in order to verify the children's understanding of the task. The participants were allowed to interrupt the experiment at will in order to rest. The experimental task was simple and interesting enough to ensure the full motivation and compliance of all participants.

***Analysis***

***Building the highly reproducible stratification model, stability-based relative clustering validation: reval***

In this work we used stability-based cluster analysis in the context of relative validation to determine whether it was possible to identify autism motor subtypes. This kind of analysis enables us to demonstrate that the obtained cluster solution is stable and generalizable. Thus, it overcomes the limitations imposed by more traditional approaches such as the high risk of overfitting or the lack of sharable criteria to establish the goodness of the stratification model. To carry out the analysis, we used the algorithm called *reval*[6] that we recently developed to facilitate this computational task. Practically, *reval* takes as input an independent training and validation sets and select the optimal number of clusters by looping across a range of possible number of clusters solutions (*k* = range[2:10]), inside of a repeated *n* cross-validation scheme applied, partitioning the training set repeatedly into the internal-training and internal-testing set, applying a clustering algorithm on both of them and simultaneously training a classifier in the internal-training set and use it to predict the clusters labels of the internal-testing set obtaining a performance metric (the misclassification error). *Reval* uses the average of the misclassification error values obtained across the *n* cross-validation repetitions for each of the possible *k* to select the optimal *k* defined as the one having the lowest *stability* (i.e., a measure that combine the classifier's misclassification error with the misclassification after random labeling). The general idea is that the optimal cluster solution in terms of number of clusters (*k*) is the most reproducible, hence a classifier trained on a first independent set (interval-training set) should be able to accurately classify the observation of another independent set (internal- testing set) resulting in a higher classification accuracy and a lower misclassification error. Once the optimal *k* has been identified, *reval* applies that *k* as the number of clusters to identify in the held-out test set. It then trains a classifier on the original training set and uses it to predict the held-out test set labels. This classifier accuracy is called generalization accuracy and it is a performance metric that allows for interpreting if the clustering solution is thoroughly reproducible in independent datasets. For more details about reval please refers to our prior published work[6].
